# Analyzing the Influencing Factors on Quality of Life of Older People with Swallowing Dysfunction in Nursing Homes in China

**DOI:** 10.1101/2023.02.15.23285953

**Authors:** Qianru Li, Laiyou Li, Ning Sun, Hongyin Wang, Shuangqin Chen, Hongyu Li, Shuang Yang

**Affiliations:** Ningbo College of Health Sciences, Ningbo, P.R. China; Zhejiang university mingzhou hospital, Yinzhou women &children’s hospital, Ningbo, P. R. China

**Keywords:** swallowing disorders, quality of life, function assessment, nursing care, older people

## Abstract

**Objective:** To investigate the prevalence of dysphagia among the older adult population in nursing homes and unravel the relevant factors that precipitate quality of life of the older adults with swallowing dysfunction in nursing homes.

**Methods:** Based on 102 individuals aged 50 years or older, with dysphagia, and who were admitted in community nursing home, we examined the demographic characteristic, the nutritional indices and the possible quality of life related variables through a cross-sectional study.

**Results:** The reliability and validity of the C-DHI scale were proven high. The incidence of swallowing disorder risk in the older adult population of care institutions is 90.2%, and the incidence of nutrition risk and malnutrition is 67.64%. The general linear regression results show that the standard regression coefficient of FOIS is −2.75, *P*=0.007. The standard regression coefficient of nutritional methods is −3.84, *P*=0.000, and the difference was statistically significant, indicating that the C-DHI scale was negatively correlated with FOIS and nutrition methods.

**Conclusions:** Swallowing disorders are also identified as a major problem amongst the older adult population in nursing homes, as there is a high prevalence of dysphagia among this population group. There are many factors related to the quality of life. The swallowing mechanism changes significantly as people age, even in the absence of chronic diseases. More attention to controllable influencing factors would improve the quality of life of older adults with swallowing dysfunction.

## 1. Introduction

Dysphagia refers to swallowing difficulties when eating or drinking caused by damage in the structure and/or function of mandible, lips, tongue, soft palate, throat, esophagus, and other organs ^[1].^ Dysphagia can occur as difficulty in chewing, saliva or food flowing out of the mouth, food staying in the mouth for a long time without swallowing, food or water flowing out of the nasal cavity (nasal reflux), choking when eating or drinking water, changes in eating habits, hoarseness, recurrent pneumonia, unexplained fever, weight loss, etc. A variety of diseases can lead to swallowing disorders, including central nervous system diseases, cranial neuropathy, neuromuscular junction diseases, oropharyngeal organic diseases, digestive system diseases, respiratory system diseases, and others. Such diseases, together with age, may lead to decreased reflexes, hypoesthesia, weakened gastrointestinal peristalsis, and loss of body position regulation, which may cause swallowing dysfunction. The swallowing mechanism changes significantly as people age, even in the absence of chronic diseases. Dysphagia can cause various complications, such as pneumonia, dehydration, malnutrition, and others, which might extend the hospital stay of patients, aggravate the disease outcome, seriously affect the quality of life (QoL) of patients, and even lead to death ^[2]^. The white paper issued by the European Association for Dysphagia has included dysphagia as one of the manifestations of the geriatric syndromes ^[3]^. The disorder is also considered to be a major problem among older adults living in nursing homes. The clinical medical staff should pay attention to that problem and strengthen the management of dysphagia in nursing care institutions.

### 1.1 Management of Dysphagia

The consensus of experts on the evaluation and treatment of swallowing disorders in China (2017 Edition) emphasizes the comprehensive management of patients with swallowing disorders in the mode of team cooperation, including the screening and evaluation, treatment, and nursing of swallowing disorders. The purpose of screening is to preliminarily understand whether there is swallowing disorder and its degree, and to find out the high-risk population. The screening should be performed routinely for patients suspecting swallowing problems, as well as for older adults and special groups, such as stroke patients, tracheotomy patients, older adults suffering frailty, among others. The safety and effectiveness of swallowing were assessed by professionals who evaluated comprehensive medical history, orofacial function, laryngeal function, and eating of the patients. Treatment includes nutrition management, function promotion, compensatory methods, and surgery.

The swallowing disorder of the older adults in care institutions is affected by numerous factors and complex causes. Although there is no treatment process and industry standard for this specific population group for swallowing disorder and its complications in China, there are expert consensus, such as China’s Expert Consensus on Assessment and Treatment of Swallowing Disorder, and China’s Expert Consensus on Rehabilitation Nursing and Care of Swallowing Disorder in Communities. Experts advocate that the older adult population, who are at risk of swallowing disorders, should be screened for as soon as possible. Therefore, care institutions for older adults should carry out screening training for swallowing disorders, improve the health management and service level for caregivers with swallowing disorders, and incorporate the regular screening of dysphagia into their daily work. The management teams in care institutions for older adults should focus on monitoring the risk population and strengthening the health management of older adults with advanced age, dementia, in healthy daily habits, stroke history and other basic diseases. Early detection, early prevention and early intervention are important measures for swallowing disorder health management.

The results of China’s seventh national population census show that the population aged 60 and above in China is 264.02 million, accounting for 18.70% of the total population. Further, the population aged 65 and above is 190.64 million, accounting for 13.50% ^[4]^, and the incidence of dysphagia among older adults aged 60 and above is 13% ^[5].^ Research shows that the incidence of dysphagia among older adults in care institutions is about 30% ^[6]^. The study found that with the increase of age, the incidence of dysphagia also increased, with incidence of 30–55% in hospitalized patients, 32–70% in Parkinson’s disease patients, and 50–75% in Alzheimer’s disease patients ^[7−8]^.

The swallowing disorder of the older adult population in care institutions deserves attention, as it can significantly reduce the QoL of the that population group. Several studies, including Lai Xiaoxing ^[9]^, show that the overall QoL of older patients with swallowing disorder is low, with total SWAL-QoL score of (45.15 ± 13.31). The dimensions evaluated, from low to high scores, include appetite, social interaction, swallowing symptoms, mental health, sleep, eating fear, eating time, fatigue, psychological burden, and language communication. Zhang Chunhua ^[10]^ investigated dysphagia in patients with stroke history in the community, reporting that the stage of stroke, degree of dysphagia, age, and eating style were the main factors affecting the patients’ QoL. The same study indicated that the QoL of patients with ischemic stroke, severe dysphagia, and nasal feeding was low, while swallowing training was the key factor for improving it. Scholars agree that good health, early detection, early prevention, and early intervention are protective factors.

The Dysphagia Index (DHI) is easy to complete, clinically available, and reliable patient independent reporting tool for older adults, used to evaluate the obstacles and inconvenience caused by the disorder to the emotional, functional, and physical aspects of personal life ^[11]^. The English version was officially released by Silbergleit in 2012 ^[12]^, and its reliability and validity have been verified multiple times ^[13−14]^. However, large samples of self-examination of dysphagia in China are limited. The C-DHI (Chinese version of the DHI) scale was first proposed by the Clinical Nutrition Department of West China Hospital of Sichuan University. After the English version was translated into Chinese, experts in clinical nutrition, geriatrics, rehabilitation and other disciplines were organized for consultation and revision ^[15]^.

In this study, several care institutions for older adults in the Zhejiang Province were selected to investigate the prevalence of swallowing disorder among older adults by a questionnaire, before analyzing the influencing factors of swallowing disorder in said care institutions in the Zhejiang Province and providing reference for the standardization of the health management of the disorder in such institutions.

## 2. Materials and Methods

### 2.1 Design

A predictive, cross-sectional design was adopted to analyze the influencing factors on the QoL of older adults with swallowing dysfunction in nursing homes, using questionnaires for data collection.

### 2.2 Participants

The participants were randomly selected from two public care institutions for older adults in Ningbo, Zhejiang Province, by cluster random sampling method and random number table method. The inclusion criteria of the respondents were as follows: 1) 50 years old or older; 2) the duration of residence in the care institution exceeds three months; 3) willing to participate in this study. Exclusion criteria were: 1) unable to communicate due to local voice, illiterate, or had disabilities in communication; 2) have severe mental or cognitive impairment. The sample consisted of 102 participants, all of which participated in the study voluntarily and were able to answer questions clearly.

### 2.3 Data Collection

#### 2.3.1 Sociodemographic Characteristics

The self-designed general situation questionnaire was used to investigate the general situation of the participants, including age, gender, education level, marital status, occupation, monthly income, and length of their residence in the care institution. Health information, such as disease diagnosis, was obtained from the medical records of the participants.

#### 2.3.2 Nutritional Status Assessment

The same participants were evaluated twice with Mini-Nutritional Assessment (MNA) at one-week intervals by trained professionals. MNA consists of 4 dimensions and 18 items in total, with a total score of 0–30. MNA score ≥24 points indicate that the nutritional status is good; 17–24 points represent a risk of malnutrition; <17 points indicate malnutrition.

#### 2.3.3 Evaluation of Dysphagia

The Functional Oral Intake Scale, FOIS, was designed in 2005 by CRARY [16], a professor of language pathology at the Swallowing Research Laboratory of the University of Florida in the United States. The scale divides the feeding function of dysphagia patients after stroke into seven grades, namely Grade 1: completely eating without mouth; Level 2: tube feeding dependence, rarely trying to eat ordinary food and liquid food; Level 3: tube feeding dependence, continuous oral intake of ordinary food and liquid food; Grade 4: eating food with single viscosity completely by mouth; Level 5: take food with various viscosity completely by mouth, but special preparation or supplement is required; Grade 6: eating a variety of viscous foods completely by mouth without special preparation, but with special food restrictions; Level 7: complete oral feeding without any restriction. FOIS assesses the oral feeding function of patients by recording the types of food they eat. Grades 1 to 3 are for patients with gastric tube indwelling, and grades 4 to 7 are for ones without gastric tube indwelling.

#### 2.3.4 Activities of Daily Living Assessment

The Barthel index was used to evaluate the daily activities of older adults. The scale includes ten items, including eating, bathing, dressing, stool control, urination control, toilet use, bed chair transfer, walking on the ground, and going up and down stairs ^[17].^ Bathing and decoration are divided into two grades (0 and 5 points); the six items of eating, dressing, stool control, urination control, toilet use, and going up and down stairs are divided into three grades (0, 5, 10 points); the two items of bed chair transfer and flat walking are divided into four grades (0, 5, 10, 15 points). All grades together give a full score of 100 points. A score of ≥ 60 points indicates that the patient has slight dysfunction, can independently complete some daily activities, but requires some help for others; 59–41 indicates moderate dysfunction and requires great help to complete daily activities; ≤ 40 points indicates severe dysfunction.

#### 2.3.5 Measures for Quality of Life

The Dysphagia Index (DHI) is one of the indexes applicable to the older adult population self-directed patient reporting that is easy to complete, clinically available, and statistically reliable. QoL was assessed by the dysphagia handicap index scale (DHI), which was first developed by Silbergleit ^[18]^. We used an adapted version of the Chinese translation of the scale, tested by the West China Hospital, Sichuan University (2022). After the translation of the Chinese version of the organization of clinical nutrition, geriatric, rehabilitation and other disciplinary experts. The scale includes 3 dimensions: Physical (items 1p–9p), Functional (items 1f–9f), and Emotional (items 1e–7e). The scale consists of 25 items to be rated using a 4-point rating scale from 0 (Never) to 4 (Always). The total score is calculated by summing all item scores, with lower scores representing higher levels of QoL.

### 2.4 Ethical Considerations

This study and the data collection instruments were approved by the Ethics Committee of the Ningbo College of Health Science. All participants provided written informed consent prior to participating in the study and were informed that participation was completely voluntary and withdrawal from the study was possible at any time.

### 2.5 Data Analysis

Data were analyzed using the SPSS 26.0 software (SPSS Inc., Chicago, IL). Descriptive statistics (frequencies, percentages, means, SD) were used to address the research questions regarding the levels of health knowledge and self-care agency. Pearson’s correlation was applied to examine the relationship between health knowledge and self-care agency. Finally, stepwise multiple regression analysis was used to examine the influence of the demographic variables and health knowledge on self-care agency.

## 3. Results

### 3.1 Participant Characteristics

From June to December 2022, a total of 102 older adults residing in nursing care institutions in Ningbo completed the survey. The overall response rate was 97.5%. The participants were predominantly females (67,64%) with an average age of 82.87±10.76 years old. Most of them had received education for ≤ 12 years (98.03%). Among them, 32 had diabetes, 62 had hypertension, 16 had coronary heart disease, 28 had senile dementia, and 32 had tumor. In addition, 2 cases were fed with nasogastric tube while the other 100 were fed orally, and 30 of them could only eat liquid diet. The mean ADL score was 47.97±39.65, with 57 patients having a poor score lower than 60. Regarding the MNA analysis among the 102 patients, 33 (32.35%) had normal nutritional status, 34 (33.33%) had nutritional risk, and 35 (34.31%) had malnutrition. The calf circumference of the patients is (25.72±5.13) cm. The analysis of swallowing function showed that the prevalence of dysphagia in this population group was 90.2%. 29% were diagnosed with mild dysphagia, 40% with moderate dysphagia, and 31% with severe dysphagia. Depending on food consistency, dysphagia to solid food was present in 60.5% of the patients, to semisolid food in 24.2%, and to liquid food in 15.3%. Nearly 53% of patients had to swallow more than once to pass the entire bolus. Table 1 shows the demographic characteristics of the participants.

**Table 1.**
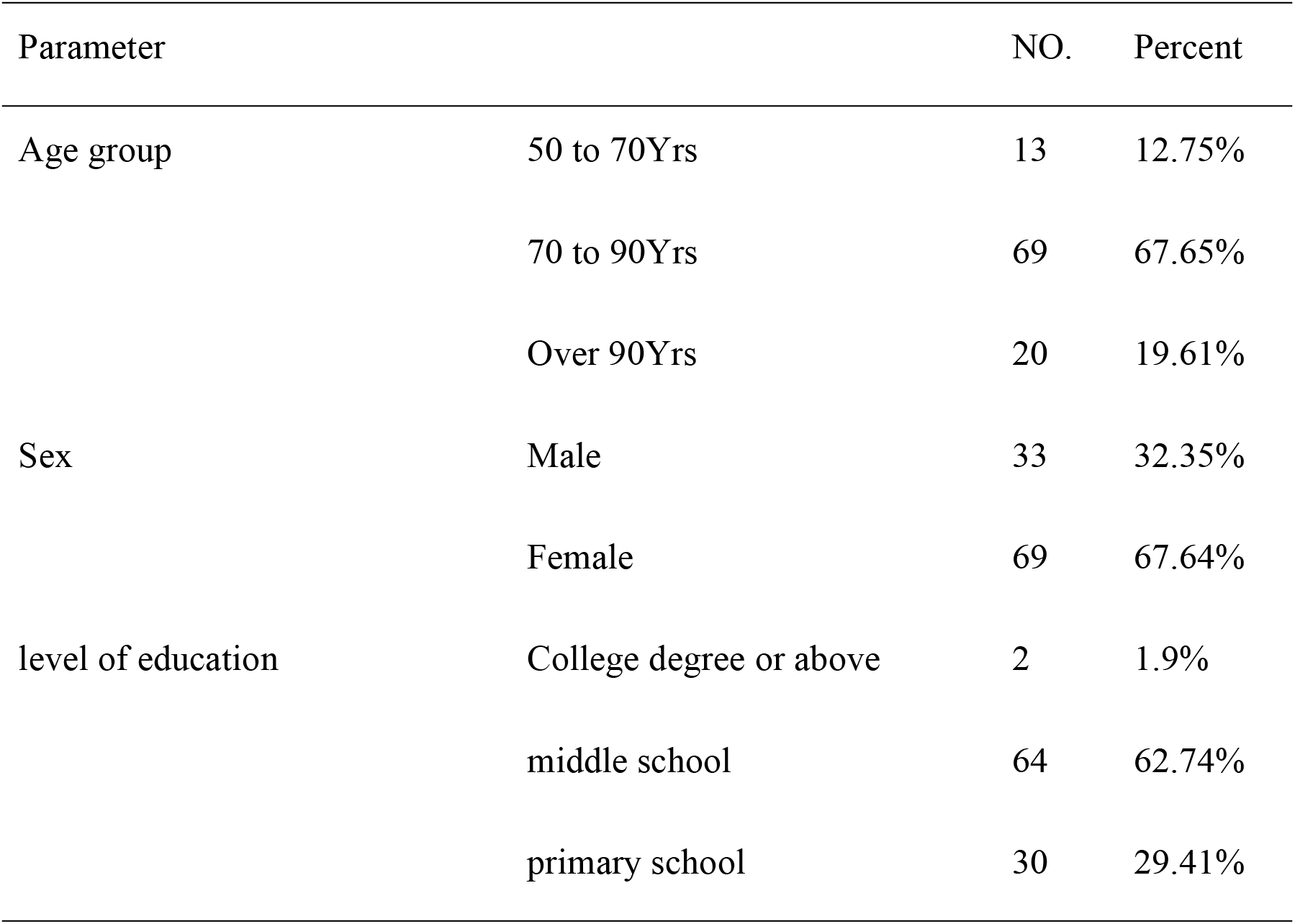
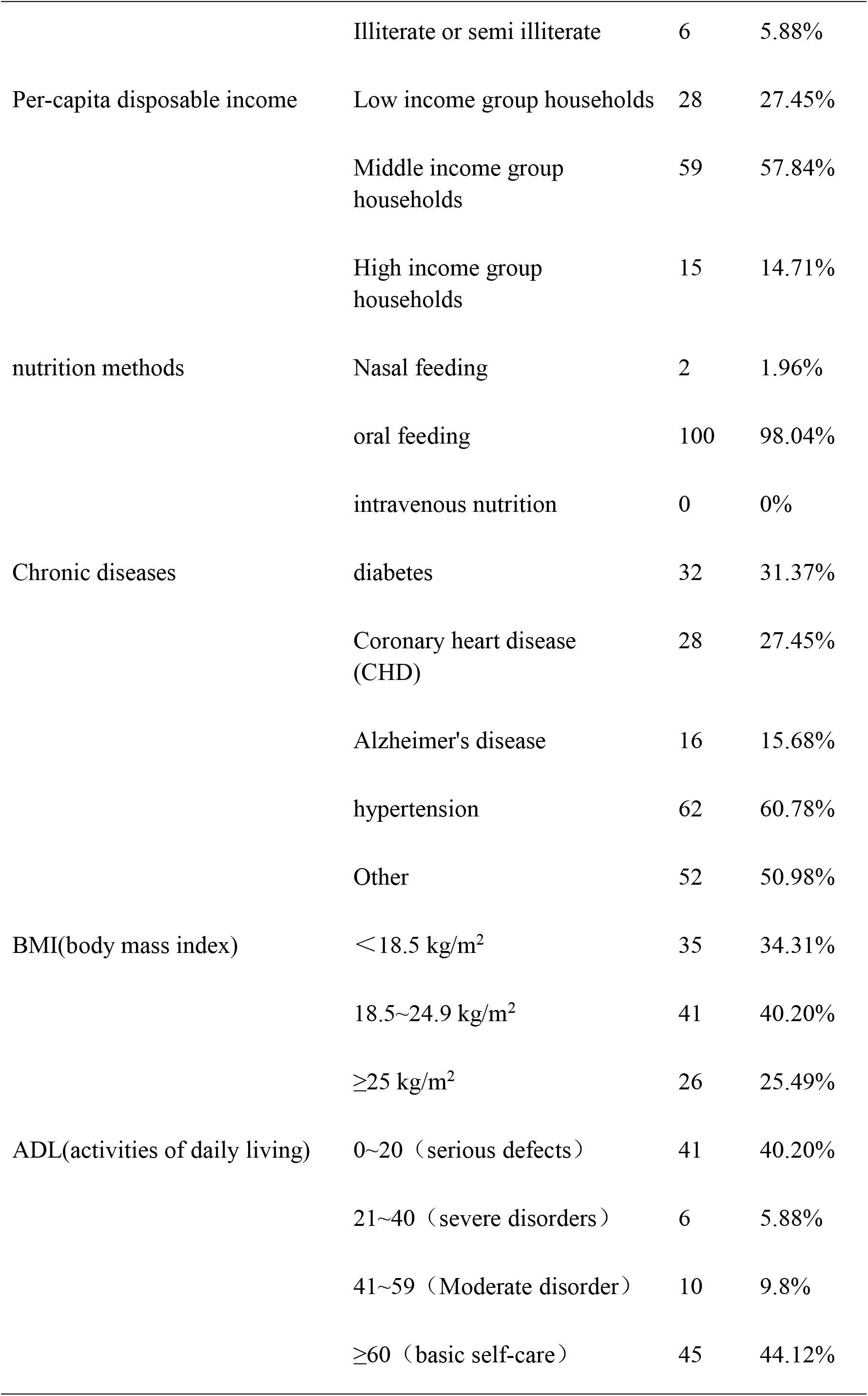
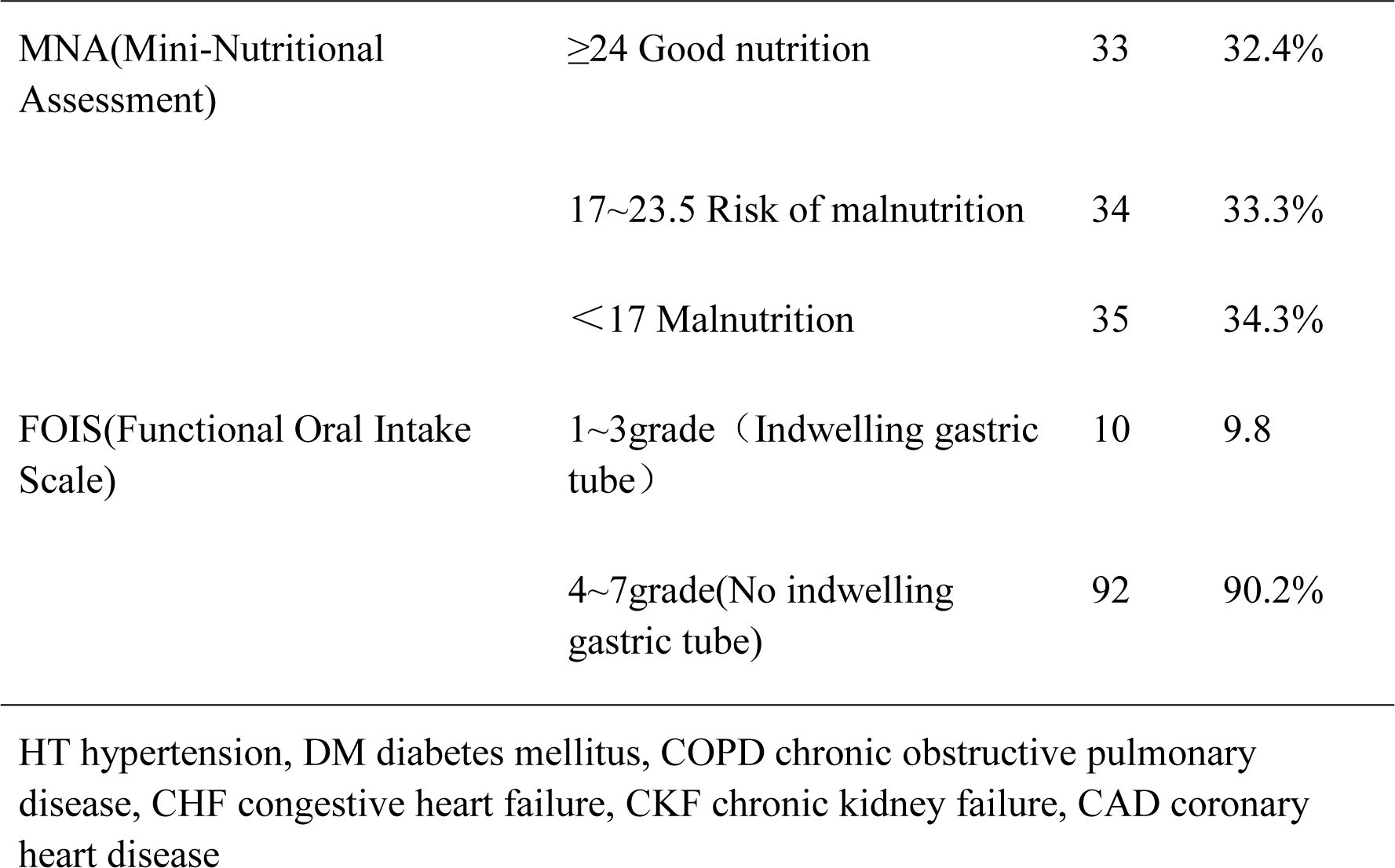
The old people’s socio-demographic characteristics

| Parameter |  | NO. | Percent |
| --- | --- | --- | --- |
| Age group | 50 to 70Yrs | 13 | 12.75% |
|  | 70 to 90Yrs | 69 | 67.65% |
|  | Over 90Yrs | 20 | 19.61% |
| Sex | Male | 33 | 32.35% |
|  | Female | 69 | 67.64% |
| level of education | College degree or above | 2 | 1.9% |
|  | middle school | 64 | 62.74% |
|  | primary school | 30 | 29.41% |
|  | Illiterate or semi illiterate | 6 | 5.88% |
| Per-capita disposable income | Low income group households | 28 | 27.45% |
|  | Middle income group households | 59 | 57.84% |
|  | High income group households | 15 | 14.71% |
| nutrition methods | Nasal feeding | 2 | 1.96% |
|  | oral feeding | 100 | 98.04% |
|  | intravenous nutrition | 0 | 0% |
| Chronic diseases | diabetes | 32 | 31.37% |
|  | Coronary heart disease (CHD) | 28 | 27.45% |
|  | Alzheimer's disease | 16 | 15.68% |
|  | hypertension | 62 | 60.78% |
|  | Other | 52 | 50.98% |
| BMI(body mass index) | <18.5 kg/m<sup>2</sup> | 35 | 34.31% |
|  | 18.5~24.9 kg/m<sup>2</sup> | 41 | 40.20% |
|  | ≥25 kg/m<sup>2</sup> | 26 | 25.49% |
| ADL(activities of daily living) | 0~20 (serious defects) | 41 | 40.20% |
|  | 21~40 (severe disorders) | 6 | 5.88% |
|  | 41~59 (Moderate disorder) | 10 | 9.8% |
|  | ≥60 (basic self-care) | 45 | 44.12% |
| MNA(Mini-Nutritional Assessment) | ≥24 Good nutrition | 33 | 32.4% |
|  | 17~23.5 Risk of malnutrition | 34 | 33.3% |
|  | <17 Malnutrition | 35 | 34.3% |
| FOIS(Functional Oral Intake Scale) | 1~3grade (Indwelling gastric tube) | 10 | 9.8 |
|  | 4~7grade(No indwelling gastric tube) | 92 | 90.2% |

HT hypertension, DM diabetes mellitus, COPD chronic obstructive pulmonary disease, CHF congestive heart failure, CKF chronic kidney failure, CAD coronary heart disease

### 3.2 Descriptive Statistics for QoL

The average score of the C-DHI was 3.76±1.48 out of 6, which was slightly higher. The average QoL score was borderline, at 61.94±13.75 out of 150. All 102 patients who were surveyed were at risk of swallowing disorder, meaning that physical factors were proven to affect the swallowing function. The median score of the functional scale was 32(18-54), and 42 cases (41.2%) were >32, indicating that the body factors also affected the swallowing function. The emotion scale had a median score of 26 (14-42), and 55 cases (53.9%) were >26, which proved that the swallowing function was also affected by emotional factors. QoL was impaired because of swallowing problems in 83.7% of the patients. Nearly 62% of them avoided eating with other persons and approximately 38% of the patients felt embarrassed at mealtimes. Patients with dysphagia presented a higher impairment in their QoL than those without swallowing problems (*P*≤0.05). The scores of each item of C-CHI scale are shown in Table 2.

**Table 2.**
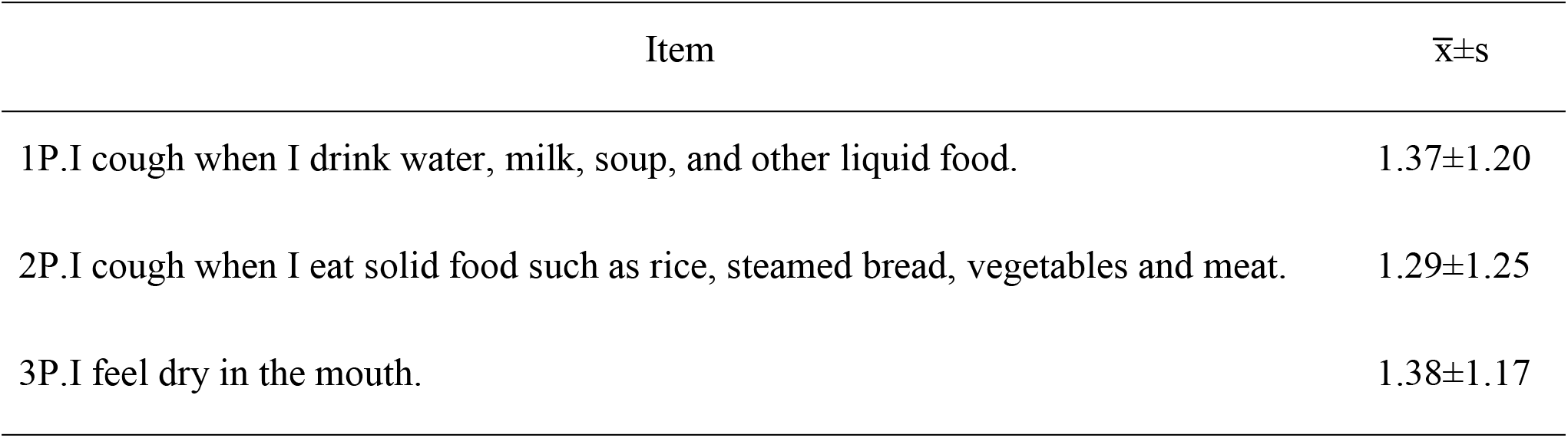
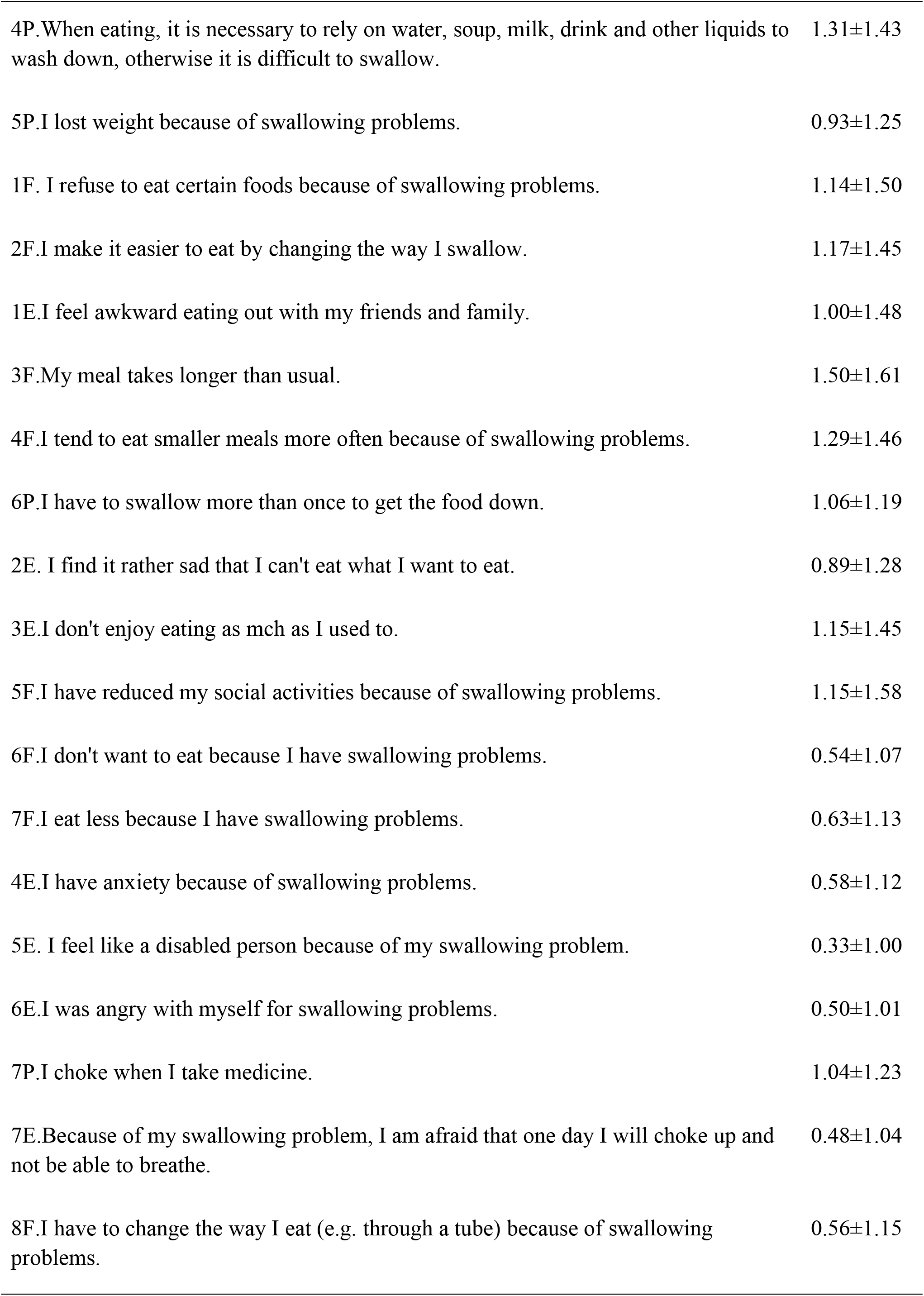
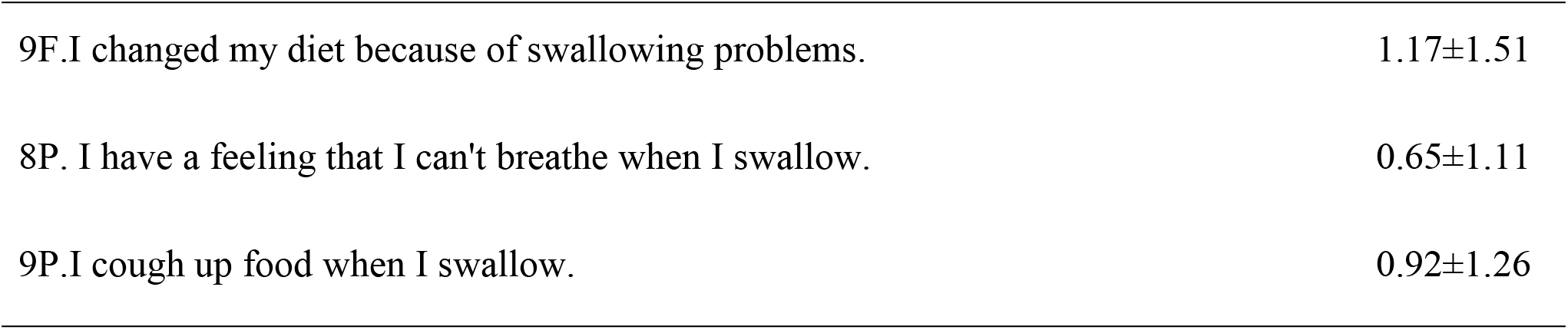
Scores of each item of C-CHI scale

| Item | $\bar{x} \pm s$ |
| --- | --- |
| 1P.I cough when I drink water, milk, soup, and other liquid food. | $1.37 \pm 1.20$ |
| 2P.I cough when I eat solid food such as rice, steamed bread, vegetables and meat. | $1.29 \pm 1.25$ |
| 3P.I feel dry in the mouth. | $1.38 \pm 1.17$ |
| 4P. When eating, it is necessary to rely on water, soup, milk, drink and other liquids to wash down, otherwise it is difficult to swallow. | 1.31±1.43 |
| 5P. I lost weight because of swallowing problems. | 0.93±1.25 |
| 1F. I refuse to eat certain foods because of swallowing problems. | 1.14±1.50 |
| 2F. I make it easier to eat by changing the way I swallow. | 1.17±1.45 |
| 1E. I feel awkward eating out with my friends and family. | 1.00±1.48 |
| 3F. My meal takes longer than usual. | 1.50±1.61 |
| 4F. I tend to eat smaller meals more often because of swallowing problems. | 1.29±1.46 |
| 6P. I have to swallow more than once to get the food down. | 1.06±1.19 |
| 2E. I find it rather sad that I can't eat what I want to eat. | 0.89±1.28 |
| 3E. I don't enjoy eating as much as I used to. | 1.15±1.45 |
| 5F. I have reduced my social activities because of swallowing problems. | 1.15±1.58 |
| 6F. I don't want to eat because I have swallowing problems. | 0.54±1.07 |
| 7F. I eat less because I have swallowing problems. | 0.63±1.13 |
| 4E. I have anxiety because of swallowing problems. | 0.58±1.12 |
| 5E. I feel like a disabled person because of my swallowing problem. | 0.33±1.00 |
| 6E. I was angry with myself for swallowing problems. | 0.50±1.01 |
| 7P. I choke when I take medicine. | 1.04±1.23 |
| 7E. Because of my swallowing problem, I am afraid that one day I will choke up and not be able to breathe. | 0.48±1.04 |
| 8F. I have to change the way I eat (e.g. through a tube) because of swallowing problems. | 0.56±1.15 |
| 9F.I changed my diet because of swallowing problems. | 1.17±1.51 |
| 8P. I have a feeling that I can't breathe when I swallow. | 0.65±1.11 |
| 9P.I cough up food when I swallow. | 0.92±1.26 |

### 3.3 General linearity was used to analyze the influencing factors of QoL in dysphagia

Regression and generalized linear model were used to analyze the influencing factors of QoL in dysphagia. Statistical analysis, the number of correlation system r=0.846, R^2^=0. 716, F=25.803, *P*<0.001, and the linear model had statistical significance. Among them, FOIS and nutrition methods (*P*<0.05) were negatively correlated with the risk and severity QoL in dysphagia, while the other factors were not significantly correlated with QoL in dysphagia (*P*>0.05). The rest of the swallowing correlation analysis of obstacles is presented in Table 3.

**Table 3.** Analysis of related influencing factors of QoL in dysphagia

| Model | Denormalization coefficient |  | Standardization coefficient | *t* | *p* |
| --- | --- | --- | --- | --- | --- |
|  | B | standard error | Beta |  |  |
| (constant) | 78.265 | 21.414 |  | 3.655 | 0.000 |
| Age | .215 | .135 | .098 | 1.596 | .114 |
| Sex | -.849 | 3.141 | -.017 | -.270 | .787 |
| level of education | -2.059 | 2.000 | -.065 | -1.030 | .306 |
| nutrition methods | -20.310 | 7.375 | -.202 | -2.754 | .007 |
| MNA | -.126 | .475 | -.030 | -.266 | .791 |
| ADL | .014 | .071 | .024 | .203 | .839 |
| BMI | .152 | .522 | .019 | .290 | .772 |
| FOIS | -7.310 | 1.901 | -.498 | -3.845 | .000 |

## 4. Discussion

### 4.1 High Risk of Swallowing Disorder among the Older Adults in Nursing Care Institutions

The research shows that the proportion of older adults at risk of swallowing disorder in nursing care institutions is relatively high, which supports the research results of relevant domestic experts. Zhang Pingping ^[19]^ and others randomly selected 837 older adults living in ten nursing homes in Weifang City, adopting the volume viscosity swallowing test for the analysis. Their results showed a 44.2% incidence of swallowing disorder among older adults in nursing homes. Xue Yu ^[20]^ verified that the Chinese swallowing disorder index scale was applied to older adults for swallowing disorder risk screening, and 146 older adults were included in a grass-roots care institution. The results showed that the incidence of swallowing disorder risk among the group in this institution was 100%, and the incidence in care institutions was also high. Foreign scholar Baijens ^[21]^ reported 60% incidence of dysphagia in care institutions for older adults, while Park et al. ^[22]^ specially investigated 395 older adults in Korean nursing homes and found dysphagia in 52.7% of the cases. The research results of many domestic and foreign experts suggest that the dysphagia incidence in care institutions for older adults is high and the complications are serious. The current situation cannot be ignored, and greater attention should be paid to the problem of dysphagia in the older adult population.

### 4.2 Main Influencing Factors of QoL in Dysphagia

The results indicated that FOIS and nutrition methods (*P*<0.05) were negatively correlated with the risk and severity of QoL in dysphagia, while the other factors were not significantly correlated with QoL in dysphagia (*P*>0.05). The results insisted with other researches. There have been both local and international studies that preliminarily explore the related factors of swallowing disorder of older adults in hospitals, communities, and care institutions, confirming that age, gender, functional status, disease history, nutritional status, and food type can all affect the swallowing disorder of older adults. Zhang Pingping ^[19]^ found a significant difference in the incidence of dysphagia among older adults with different ages and health conditions (*P*<0.05). The participants with stroke history, Alzheimer’s disease, and Parkinson’s disease mainly had oral dysfunction (*P*<0.001), and the ones with chronic respiratory disease mainly had pharyngeal and airway protection dysfunction (*P*<0.01). Zhao Wenting et al. ^[23]^ examined 382 patients in the geriatric department of a Grade III A general hospital as the research objects, reporting 96 patients with dysphagia, accounting for 25.1%, as well as 89 cases (23.3%) with sarcopenia. Multivariate analysis showed that age, degree of muscular deficiency, and self-care ability were the main influencing factors of dysphagia. Wang Yue et al. ^[24]^ investigated the occurrence of swallowing dysfunction in older adults with mild cognitive impairment and analyzed the factors affecting the occurrence. Logistic regression model analysis results showed that age ≥70 years old, asthenia score ≥3 points, visual and auditory impairment, and tooth defect ≥6 were risk factors for swallowing dysfunction in older adults with mild cognitive impairment, while swallowing training and activities of daily living score ≥60 points were protective factors for the same population group (*P*<0.05).Some experts ^[25−26]^ analyzed older adults in the community, suggesting that age (≥80 years old), education level (junior high school and below), history of stroke, history of cough, score of the simple mental state scale (≤23 points), activity of daily living score (≤60 points), chewing function score (≤3 points), and number of drugs (≥3 kinds) were all risk factors related to swallowing disorder, while food type (universal food) was found to be a protective factor. Numerous studies have shown that the risk of dysphagia increases with age ^[27]^. Moreover, the weaker the ability of older adults to perform daily activities, the higher their risk for dysphagia. The statistical test of this study shows that there are differences in age and activities of daily living in dysphagia, which are statistically significant (*P*<0.05), indicating that the disorder is related to age and daily living activities.

### 4.3 Limitations of This Study

Our study was limited by its modest sample size, which may have impacted the results, as well as the width of confidence intervals. Variability in the sample may have also contributed to the width of confidence intervals. It should not be used as a single or principal measure as it is influenced by the individuals’ cognitive condition. Results pointed to a significant statistical relation between objective and subjective measures, thus indicating that a self-perception test should be included in the assessment of swallowing disorders in a nursing home population.

## 5. Conclusion

Investigating the status of QoL in dysphagia for older people is necessary to efficiently and objectively identify problems and provide early intervention in nursing homes, where dysphagia is responsible for many acute and chronic respiratory diseases, increasing the state of dependency, and contributing to rates of morbidity and mortality.There are many factors related to the QoL. The swallowing mechanism changes significantly as people age, even in the absence of chronic diseases. More attention to controllable influencing factors would improve the QoL of older adults with swallowing dysfunction.

## Data Availability

The data underlying the results presented in the study are available from care institutions

## Author Contributions

The authors were responsible for the paper as follows: QRL, conception, design, analysis, and data interpretation, drafting the manuscript, revising the manuscript, and its final approval; LYL, acquisition of data, project administration, manuscript revisions, and its final approval; HYW and SQC, formal analysis, manuscript revision, and final approval; HYL and SY, conception, manuscript revision, and final approval; and NS, conception, design, funding acquisition, project administration, manuscript revision, and final approval. All the authors have read and approved the final manuscript.

## Acknowledgments

The authors would like to thank the older adults who participated in the study. The authors also thank the source of financial grants: Zhejiang Province Universities Major Humanities and Social Sciences Tackling Plan Project and Zhejiang Philosophy and Social Science Planning Project.

